# Genal: A Python Toolkit for Genetic Risk Scoring and Mendelian Randomization

**DOI:** 10.1101/2024.05.23.24307776

**Authors:** Cyprien A. Rivier, Santiago Clocchiatti-Tuozzo, Shufan Huo, Victor Torres-Lopez, Daniela Renedo, Kevin Sheth, Guido J. Falcone, Julian N. Acosta

## Abstract

**Summary:** Here we present Genal, a python module for population genetic analyses. It includes functionalities for cleaning and formatting Single Nucleotide Polymorphism (SNP)-level data, clumping, lifting, SNP association testing, polygenic risk scoring, and mendelian randomization analyses, all within a single module. It was designed with user-friendliness in mind, aiming to reduce the programming skills threshold required for medical scientists to perform genetic epidemiology studies. Genal eliminates the need to interact with the command line and switch between multiple R packages with different requirements, making it considerably more accessible to a wide range of researchers. Genal draws on concepts from several well-established tools, ensuring that users have access to rigorous statistical techniques in the intuitive Python environment.

## Introduction

As the volume of genetic association data from Genome-Wide Association Studies (GWAS) continues to expand, methodologies such as Polygenic Risk Scores (PRS) and Mendelian Randomization (MR) analyses have rapidly gained importance within and beyond the field of genetic epidemiology. PRS synthesize the vast array of genetic variants across the genome to quantify an individual’s predisposition to various diseases and traits, offering a powerful tool for risk stratification and personalized medicine. Concurrently, MR utilizes genetic variants as instruments for causal inference between traits and clinical outcomes, enabling researchers to identify potential therapeutic targets and to understand disease etiologies without the confounding biases typical of observational studies.

Despite the increasing utility of PRS and MR in leveraging genetic association data for clinical and research purposes, their application is often hindered by the multi-step processes involved in data preparation and analysis. The need for specialized knowledge in bioinformatics and the use of disparate tools for different steps of the analytical process, each requiring preparing the data in a different way, further complicate the workflow, making it less accessible to researchers with non-computational backgrounds.

Python, trailing only behind JavaScript in popularity^1^, stands as one of the most widely used and fastest-growing programming languages globally^2^. Its ascent to prominence is largely attributable to its simplicity, readability, and its adaptability that allows for the seamless integration of statistical analyses, machine learning algorithms, and data visualization^3^. In this context, we present Genal, a python toolkit that integrates Single Nucleotide Polymorphisms (SNP)-level data handling, clumping, lifting, SNP association testing, polygenic risk scoring, and mendelian randomization functionalities into one user-friendly package. Genal eliminates the need to install multiple R packages with various dependencies and the data formatting required when switching between them. It also entirely avoids the need to interact with the command line, making it considerably more accessible to a wide range of researchers. We also compare the accuracy and performance of Genal to the tools currently used to perform these tasks.

## Implementation

### The *Geno* class

The ‘*Geno*’ class is the central element of the Genal package. It is designed to store SNP association data, and all functionalities of the toolkit are implemented as methods of this class. The main attribute of the ‘*Geno*’ class is a pandas DataFrame^4^, which stores the information for each SNP as rows and includes columns for the genomic position, reference SNP cluster ID (rsid), point estimate, standard error, and p-value of the association with the trait, effect and non-effect alleles, and the frequency of the effect allele. Not all columns must be present to create an instance of the ‘*Geno*’ class. If some of them are missing, the ‘*preprocess_data*’ method can derive it from the existing columns and genetic reference panels. For example, it can determine the genomic position from the rsid, or conversely, ascertain the rsid from the genomic position. Similarly, the standard error can be deduced from the p-value and vice versa. The ‘*preprocess_data*’ method also performs a variety of checks to clean the data, eliminating errors, missing values, or invalid entries across each column. It ensures the data is formatted correctly for use in the toolkit’s downstream methods. Users have the flexibility to customize this list of checks. The method provides feedback by displaying stepwise results, allowing users to monitor the number of SNPs filtered out at each stage.

### Clumping and risk scoring

After SNP data is loaded into a ‘*Geno*’ instance and has been cleaned and formatted, several genetic methods can be applied directly with just a single line of code. The clump method allows for clumping, returning a new Geno instance with data clumped based on specified thresholds and reference panels. To calculate a PRS for a population, the ‘*prs*’ method is used, providing the option to use proxies at certain thresholds. When dealing with data separated by chromosomes, Genal manages the extraction and merging of the required SNPs. The PRS method can be applied to any Geno instance, whether it has been processed through clumping or not. Both the clumping and PRS methods are built on top of Plink^5^, the only external dependency that needs to be installed. Reference panels used are the 1K genomes.

### Mendelian Randomization

Running Mendelian Randomization (MR) with Genal is straightforward, starting with the preparation of two ‘*Geno*’ instances. The first should include SNPs chosen as instruments and their association with the exposure trait, typically a ‘*Geno*’ instance resulting from the clumping step. The second ‘*Geno*’ instance is expected to hold summary statistics for the association with the outcome trait, which will be used to extract the values corresponding to the instruments. After formatting both Geno instances with ‘*preprocess_data*’, the necessary dataframe for MR is generated after invoking the ‘*query_outcome*’ method on the exposure ‘*Geno*’ instance, with the results stored as an attribute. In cases where not all instruments are present in the outcome data, the proxy option allows for the use of alternate SNPs within specified thresholds. To conduct MR analysis, one simply calls the *‘MR*’ method. This method takes care of SNP harmonization and deals with palindromic SNPs (the nucleotide sequence is identical when read forward and backward) as per the user’s specifications, similar to the process outlined in TwoSampleMR^6,7^. Users can specify which MR methods to use through arguments, along with advanced settings like the number of bootstrapping iterations for particular MR analyses, or options to return heterogeneity results. The outcomes of the MR analysis are displayed in a table format and can be visually represented using the *‘MR_plot*’ method. For addressing potential horizontal pleiotropy, beyond the weighted median and MR-Egger methods available within the *‘MR*’ method, the MR-PRESSO^8^ algorithm can be called via the *‘MRpresso*’ method. All the functionalities mentioned are implemented fully in Python and the bootstrapping and MR-PRESSO methods utilize parallel processing to speed up the execution.

## Example

In the supplementary material, we provide a tutorial along with an example where we construct a PRS for systolic blood pressure and examine its genetic influence on the risk of all-cause stroke using MR. This analysis leverages public GWAS summary statistics^9,10^. For the MR analyses, we achieve results that match those obtained with TwoSampleMR, the most commonly used tool for such analyses. Additionally, we present a Python-based parallel implementation of MR-PRESSO. Our results are consistent with the original R version of MR-PRESSO, and the method implemented in Genal is 87.5% faster. Specifically, for an analysis involving 1499 SNPs and 30,000 iterations of random data generation, the original R version required 15 hours and 50 minutes, whereas our Genal version, running on an AMD Ryzen 5900x CPU with 12 cores at 3.7 GHz, completed the task in 1 hour and 58 minutes.

We also used Genal to replicate analyses from previously published papers^11–14^ and abstracts^14–17^, demonstrating that the results can be obtained using only a few lines of code.

## Supporting information

Tutorial and Validation

## Data availability statement

The package is available on Pypi (https://pypi.org/project/genal-python/) and the code is openly available on Github with a tutorial: https://github.com/CypRiv/genal

## References

1. The top programming languages. The State of the Octoverse. Accessed March 14, 2024. https://octoverse.github.com/2022/top-programming-languages

2. Srinath KR. Python – The Fastest Growing Programming Language. 04(12).

3. Sharma A, Khan F, Sharma D, Gupta DS. Python: The Programming Language of Future. 2020;6(12).

4. pandas - Python Data Analysis Library. Accessed March 25, 2024. https://pandas.pydata.org/

5. Purcell S, Neale B, Todd-Brown K. PLINK: a tool set for whole-genome association and population-based linkage analyses. Am J Hum Genet. 2007;81(3):559–575. doi:10.1086/519795

6. Orienting the causal relationship between imprecisely measured traits using GWAS summary data | PLOS Genetics. Accessed March 25, 2024. https://journals.plos.org/plosgenetics/article?id=10.1371/journal.pgen.1007081

7. Hemani G, Zheng J, Elsworth B, et al. The MR-Base platform supports systematic causal inference across the human phenome. Elife. 2018;7:e34408. doi:10.7554/eLife.34408

8. Verbanck M, Chen CY, Neale B, Do R. Detection of widespread horizontal pleiotropy in causal relationships inferred from Mendelian randomization between complex traits and diseases. Nat Genet. 2018;50(5):693–698. doi:10.1038/s41588-018-0099-7

9. Evangelou E, Warren HR, Mosen-Ansorena D. Genetic analysis of over 1 million people identifies 535 new loci associated with blood pressure traits. Nat Genet. 2018;50(10):1412–1425. doi:10.1038/s41588-018-0205-x

10. Debette S, Mishra A, Malik R, et al. Stroke genetics informs drug discovery and risk prediction across ancestries. Published online January 12, 2022. doi:10.21203/rs.3.rs-1175817/v1

11. Rivier CA, Szejko N, Renedo D, et al. Polygenic Susceptibility to Hypertension and Cognitive Performance in Middle-aged Persons Without Stroke or Dementia. Neurology. 2023;101(5):e512–e521. doi:10.1212/WNL.0000000000207427

12. Rivier CA, Renedo DB, de Havenon A, et al. Association of Poor Oral Health With Neuroimaging Markers of White Matter Injury in Middle-Aged Participants in the UK Biobank. Neurology. 2024;102(2):e208010. doi:10.1212/WNL.0000000000208010

13. Rivier CA, Clocchiatti-Tuozzo S, Misra S, et al. Polygenic Risk of Epilepsy and Post-Stroke Epilepsy. Published online September 18, 2023:2023.09.18.23295739. doi:10.1101/2023.09.18.23295739

14. Demarais ZS, Conlon CJ, Huo S, et al. Polygenic Susceptibility to Diabetes and Poor Glycemic Control in Stroke Survivors. Published online September 18, 2023:2023.09.18.23295736. doi:10.1101/2023.09.18.23295736

15. Rivier C, Clocchiatti-Tuozzo S, Huo S, et al. Abstract TMP77: Oral Health as a Risk Factor for Spontaneous Intracerebral Hemorrhage: A Mendelian Randomization Analysis. Stroke. 2024;55(Suppl_1):ATMP77–ATMP77. doi:10.1161/str.55.suppl_1.TMP77

16. Huo S, Rivier C, Clocchiatti-Tuozzo S, et al. Polygenic Resistance to Blood Pressure Treatment and Stroke Risk (S6.001). Neurology. 2024;102(17_supplement_1):6042. doi:10.1212/WNL.0000000000206202

17. Rivier C, Acosta J, Renedo D, Sheth KN, Falcone GJ. Abstract 32: Neighborhood Deprivation And Polygenic Contribution To Acute Ischemic Stroke: Results From The All Of Us Research Program. Stroke. 2023;54(Suppl_1):A32–A32. doi:10.1161/str.54.suppl_1.32

