## Supplementary material for "Genal: A Python Toolkit for Genetic Risk Scoring and Mendelian Randomization": Tutorial and Validation

### Table of contents


1. Introduction
2. Requirements for the genal module
3. Installation and how to use genal
    i. Installation
4. Tutorial and presentation of the main tools
    i. Data loading
    ii. Data preprocessing
    iii. Clumping
    iv. Polygenic Risk Scoring
    v. Mendelian Randomization and validation with R
    vi. SNP-association testing
    vii. Lifting
5. Examples from published papers


#### Introduction

Genal is a python module designed to make it easy to run genetic risk scores and mendelian randomization analyses. It integrates a collection of tools that facilitate the cleaning of single nucleotide polymorphism data (usually derived from Genome-Wide Association Studies) and enable the execution of clinical population genetic workflows. The functionalities provided by genal include clumping, lifting, association testing, polygenic risk scoring, and Mendelian randomization analyses, all within a single Python module.

The module prioritizes user-friendliness and intuitive operation, aiming to reduce the complexity of data analysis for researchers. Despite its focus on simplicity, Genal does not sacrifice the depth of customization or the precision of analysis. Researchers can expect to maintain analytical rigour while benefiting from the streamlined experience.

Genal draws on concepts from well-established R packages such as TwoSampleMR, MR-Presso, MendelianRandomization, and gwasvcf, adapting their proven methodologies to the Python environment. This approach ensures that users have access to tried and tested techniques with the versatility of Python's data science tools.

#### Requirements for the genal module

***Python 3.9 or later***. https://www.python.org/

#### Installation and How to use the genal module

##### Installation

Download and install the package with pip:

```
pip install genal-python
```

And it can be imported in a python environment with:

```python
import genal
```

The Github page of genal is: Link to github

The main genal functionalities require a working installation of PLINK v1.9 that can be downloaded here: https://www.cog-genomics.org/plink/ Once downloaded, the path to the plink executable can be set with:

```
genal.set_plink(path="/path/to/plink/executable/file")
```

#### Tutorial

For this tutorial, we will obtain genetic instruments for systolic blood pressure (SBP), compute a Polygenic Risk Score (PRS), and run a Mendelian Randomization analysis to investigate the genetically-determined effect of SBP on the risk of stroke. As a validation step, we will compare our results to the ones obtained with R packages. We will utilize summary statistics from Genome-Wide Association Studies (GWAS) and individual-level data from the UK Biobank. The steps include:

- Data loading
- Data preprocessing
  - Perform checks and cleaning operations on SNP-level data for appropriate formatting
- Clump data to identify significant, independent SNPs as genetic instruments for SBP
- Build a genomic risk score for SBP in a test population
  - Include risk score calculations with proxies
- Perform Mendelian Randomization
  - Analyze SBP as an exposure and acute stroke as an outcome
  - Comparison of the results to those obtained with the TwoSampleMR package.
  - Plot the results
  - Conduct sensitivity analyses using the weighted median, MR-Egger, and MR-PRESSO methods
  - Comparison of the results to those obtained with the TwoSampleMR and MR-PRESSO packages.
- Calibrate SNP-trait weights with individual-level genetic data
  - Execute single-SNP association tests for calibrating SBP genetic instruments
- Utility tools demonstration
  - Data lifting to another genomic build
    - In pure Python
    - Using LiftOver
  - Phenoscanner (to be added)

##### Data loading

We begin with publicly available summary statistics from a large GWAS study of systolic blood pressure. Link to study. After downloading and unzipping the summary statistics, we load them into a pandas DataFrame:

```
import pandas as pd
sbp_gwas = pd.read_csv("Evangelou_30224653_SBP.txt", sep=" ")
sbp_gwas.head(5)
```

| MarkerName | Allele1 | Allele2 | Freq1 | Effect | StdErr | P | TotalSampleSize | N_effective |
|---|---|---|---|---|---|---|---|---|
| 10:100000625:SNP | a | g | 0.5660 | 0.0523 | 0.0303 | 0.083940 | 738170 | 736847 |
| 10:100000645:SNP | a | c | 0.7936 | 0.0200 | 0.0372 | 0.591100 | 738168 | 735018 |
| 10:100003242:SNP | t | g | 0.8831 | 0.1417 | 0.0469 | 0.002526 | 738168 | 733070 |
| 10:100003304:SNP | a | g | 0.9609 | 0.0245 | 0.0838 | 0.769800 | 737054 | 663809 |
| 10:100003785:SNP | t | c | 0.6406 | -0.0680 | 0.0313 | 0.029870 | 738169 | 735681 |

We can now load this data into a `genal.Geno` instance. The `genal.Geno` class is the central piece of the package. It is designed to store Single Nucleotide Polymorphisms (SNP) data and make it easy to preprocess and clean.

The `genal.Geno` takes as input a pandas dataframe where each row corresponds to a SNP, with columns describing the position and possibly the effect of the SNP for the given trait (SBP in our case). To indicate the names of the columns, the following arguments can be passed:

- **CHR**: Column name for chromosome. Defaults to `'CHR'`.
- **POS**: Column name for genomic position. Defaults to `'POS'`.

- **SNP**: Column name for SNP identifier (rsid). Defaults to `'SNP'` .
- **EA**: Column name for effect allele. Defaults to `'EA'` .
- **NEA**: Column name for non-effect allele. Defaults to `'NEA'` .
- **BETA**: Column name for effect estimate. Defaults to `'BETA'` .
- **SE**: Column name for effect standard error. Defaults to `'SE'` .
- **P**: Column name for effect p-value. Defaults to `'P'` .
- **EAF**: Column name for effect allele frequency. Defaults to `'EAF'` .

After inspecting the dataframe, we first need to extract the chromosome and position information from the `MarkerName` column into two new columns `CHR` and `POS` :

```
sbp_gwas[["CHR", "POS", "Filler"]] = sbp_gwas["MarkerName"].str.split(":", expand=True)
sbp_gwas.head(5)
```

| MarkerName | Allele1 | Allele2 | Freq1 | Effect | StdErr | P | TotalSampleSize | N_effective | CHR |
|---|---|---|---|---|---|---|---|---|---|
| 10:100000625:SNP | a | g | 0.5660 | 0.0523 | 0.0303 | 0.083940 | 738170 | 736847 | 10 |
| 10:100000645:SNP | a | c | 0.7936 | 0.0200 | 0.0372 | 0.591100 | 738168 | 735018 | 10 |
| 10:100003242:SNP | t | g | 0.8831 | 0.1417 | 0.0469 | 0.002526 | 738168 | 733070 | 10 |
| 10:100003304:SNP | a | g | 0.9609 | 0.0245 | 0.0838 | 0.769800 | 737054 | 663809 | 10 |
| 10:100003785:SNP | t | c | 0.6406 | -0.0680 | 0.0313 | 0.029870 | 738169 | 735681 | 10 |

And it can now be loaded into a `genal.Geno` instance:

```
import genal
SBP_Geno = genal.Geno(sbp_gwas, CHR="CHR", POS="POS", EA="Allele1", NEA="Allele2", BETA="Effect",
SE="StdErr", P="P", EAF="Freq1", keep_columns=False)
```

The last argument ( `keep_columns = False` ) indicates that we do not wish to keep the other (non-main) columns in the dataframe. Defaults to `True` .

> **Note:**
>
> Make sure to read the readme file usually provided with the summary statistics to identify the correct columns. It is particularly important to correctly identify the allele that represents the effect allele. Also, you do not need all columns to move forward, as some can be inputted as we will see next.

#### Data preprocessing

Now that we have loaded the data into a `genal.Geno` instance, we can begin cleaning and formatting it. Methods such as Polygenic Risk Scoring or Mendelian Randomization require the SNP data to be in a specific format. Also, raw summary statistics can sometimes contain missing or invalid values that need to be handled. Additionally, some columns may be missing from the data (such as the SNP rsid column, or the non-effect allele column) and these columns can be created based on existing ones and a reference panel.

Genal can run all the basic cleaning and preprocessing steps in one command:

```
SBP_Geno.preprocess_data(preprocessing = 'Fill_delete')
```

The `preprocessing` argument specifies the global level of preprocessing applied to the data:

- `preprocessing = 'None'` : The data won't be modified.
- `preprocessing = 'Fill'` : Missing columns will be added based on reference data and invalid values set to NaN, but no rows will be deleted.
- `preprocessing = 'Fill_delete'` : Missing columns will be added, and all rows containing missing, duplicated, or invalid values will be deleted. This option is recommended before running genetic methods. Defaults to `'Fill'` .

By default, and depending on the global preprocessing level ( `'None'` , `'Fill'` , `'Fill_delete'` ) chosen, the `preprocess_data` method of `genal.Geno` will run the following checks:

- Ensure the `CHR` (chromosome) and `POS` (genomic position) columns are integers.

- Ensure the `EA` (effect allele) and `NEA` (non-effect allele) columns are uppercase characters containing A, T, C, G letters. Multiallelic values are set to NaN.
- Validate the `P` (p-value) column for proper values.
- Check for no duplicated SNPs based on rsid.
- Determine if the `BETA` (effect) column contains beta estimates or odds ratios, and log-transform odds ratios if necessary.
- Create `SNP` column using a reference panel if CHR and POS columns are present.
- Create `CHR` and/or `POS` column using a reference panel if `SNP` column is present.
- Create `NEA` (non-effect allele) column using a reference panel if `EA` (effect allele) column is present.
- Create the `SE` (standard-error) column if the `BETA` and `P` (p-value) columns are present.
- Create the `P` column if the `BETA` and `SE` columns are present.

If you do not wish to run certain steps, or wish to run only certain steps, you can use additional arguments. For more information, please refer to the `genal.Geno.preprocess_data` method in the API documentation.

In our case, the `SNP` column (for SNP identifier - rsid) was missing from our dataframe and has been added based on a 1000 genome reference panel:

```
Using the EUR reference panel.
The SNP column (rsID) has been created. 197511(2.787%) SNPs were not found in the reference data and their ID set to CHR:POS:EA.
The BETA column looks like Beta estimates. Use effect_column='OR' if it is a column of Odds Ratios.
```

You can always check the data of a `genal.Geno` instance by accessing the `data` attribute:

```
SBP_Geno.data
```

| | EA | NEA | EAF | BETA | SE | P | CHR | POS | SNP |
|---|---|---|---|---|---|---|---|---|---|
| 0 | A | G | 0.5660 | 0.0523 | 0.0303 | 0.083940 | 10 | 100000625 | rs7899632 |
| 1 | A | C | 0.7936 | 0.0200 | 0.0372 | 0.591100 | 10 | 100000645 | rs61875309 |
| 2 | T | G | 0.8831 | 0.1417 | 0.0469 | 0.002526 | 10 | 100003242 | rs12258651 |
| 3 | A | G | 0.9609 | 0.0245 | 0.0838 | 0.769800 | 10 | 100003304 | rs72828461 |
| 4 | T | C | 0.6406 | -0.0680 | 0.0313 | 0.029870 | 10 | 100003785 | rs1359508 |
| ... | .. | .. | ... | ... | ... | ... | ... | ... | ... |
| 7088120 | A | G | 0.9028 | -0.0184 | 0.0517 | 0.722300 | 9 | 99999468 | rs10981301 |

And we see that the `SNP` column with the rsids has been added based on the reference data. You do not need to obtain the 1000 genome reference panel yourself, genal will download it the first time you use it. By default, the reference panel used is the european (eur) one. You can specify another valid reference panel (afr, eas, sas, amr) with the reference_panel argument:

```
SBP_Geno.preprocess_data(preprocessing = 'Fill_delete', reference_panel = "afr")
```

You can also use a custom reference panel by specifying to the reference_panel argument a path to bed/bim/fam files (without the extension).

#### Clumping

Clumping is the step at which we select the SNPs that will be used as our genetic instruments in future Polygenic Risk Scores and Mendelian Randomization analyses. The process involves identifying the SNPs that are strongly associated with our trait of interest (systolic blood pressure in this tutorial) and are independent from each other. This second step ensures that selected SNPs are not highly correlated, (i.e., they are not in high linkage disequilibrium). For this step, we again need to use a reference panel.

The SNP-data loaded in a `genal.Geno` instance can be clumped using the `genal.Geno.clump` method. It will return another `genal.Geno` instance containing only the clumped data:

```
SBP_clumped = SBP_Geno.clump(p1 = 5e-8, r2 = 0.1, kb = 250, reference_panel = "eur")
```

It will output the number of instruments obtained::

```
Using the EUR reference panel.
Warning: 760  top variant IDs missing
1545 clumps formed from 73594 top variants.
```

You can specify the thresholds you want to use for the clumping with the following arguments:

- `p1` : P-value threshold during clumping. SNPs with a P-value higher than this value are excluded. Defaults to `5e-8` .
- `r2` : Linkage disequilibrium threshold for the independence check. Takes values between 0 and 1. Defaults to `0.1` .
- `kb` : Genomic window used for the independence check (the unit is thousands of base-pair positions). Defaults to `250` .
- `reference_panel` : The reference population used to derive linkage disequilibrium values and select independent SNPs. Defaults to `eur` .

#### Polygenic Risk Scoring

Computing a Polygenic Risk Score (PRS) can be done in one line with the `genal.Geno.prs` method:

```
SBP_clumped.prs(name = "SBP_prs", path = "path/to/genetic/files")
```

The genetic files of the target population can be either contained in one triple of bed/bim/fam files with information for all SNPs, or divided by chromosome (one bed/bim/fam triple for chr 1, another for chr 2, etc...). In the latter case, provide the path by replacing the chromosome number by `$` and genal will extract the necessary SNPs from each chromosome and merge them before running the PRS. For instance, if the genetic files are named `Pop_chr1.bed` , `Pop_chr1.bim` , `Pop_chr1.fam` , `Pop_chr2.bed` , ..., you can use:

```
SBP_clumped.prs(name = "SBP_prs", path = "Pop_chr$")
```

The `name` argument specifies the name of the .csv file that will be saved with the individual risk scores. The output of the `genal.Geno.prs` method will include how many SNPs were used to compute the risk score. It can happen that some of the SNPs are multiallelic in the genetic data (even if they are not multiallelic in our SNP data) and need to be excluded. It can also happen that some of the SNPs are missing from the genetic files of the target population (for instance if the data has not been imputed):

```
CHR/POS columns present: SNPs searched based on genomic positions.
Extracting SNPs for each chromosome...
SNPs extracted for chr1.
SNPs extracted for chr2.
SNPs extracted for chr3.
SNPs extracted for chr4.
SNPs extracted for chr5.
SNPs extracted for chr6.
SNPs extracted for chr7.
SNPs extracted for chr8.
SNPs extracted for chr9.
SNPs extracted for chr10.
SNPs extracted for chr11.
SNPs extracted for chr12.
SNPs extracted for chr13.
SNPs extracted for chr14.
SNPs extracted for chr15.
SNPs extracted for chr16.
SNPs extracted for chr17.
SNPs extracted for chr18.
SNPs extracted for chr19.
SNPs extracted for chr20.
SNPs extracted for chr21.
SNPs extracted for chr22.
Merging SNPs extracted from each chromosome...
Created bed/bim/fam fileset with extracted SNPs: tmp_GENAL/4f4ce6a7_allchr
Extraction completed. 786(50.874%) SNPs were not extracted from the genetic data.
Computing a weighted PRS using tmp_GENAL/4f4ce6a7_allchr data.
The PRS computation was successful and used 759/1545 (49.126%) SNPs.
PRS data saved to SBP_prs.csv
```

Here, we see that about half of the SNPs were not extracted from the data. In such cases, we may want to try and salvage some of these SNPs by looking for proxies (SNPs in high linkage disequilibrium, i.e. highly correlated SNPs). This can be done by specifying the `proxy = True` argument:

```python
SBP_clumped.prs(name = "SBP_prs" ,path = "Pop_chr$", proxy = True, reference_panel = "eur", r2=0.8,
kb=5000, window_snps=5000)
```

and the output is:

```
CHR/POS columns present: SNPs searched based on genomic positions.
Identifying the SNPs present in the genetic data...
759 SNPs out of 1545 are present in the genetic data.
Searching proxies for 786 SNPs...
Using the EUR reference panel.
Filtering the potential proxies with the searchspace provided.
Found proxies for 578 missing SNPs.
7(0.455%) duplicated SNPs have been removed. Use keep_dups=True to keep them.
Extracting SNPs for each chromosome...
SNPs extracted for chr1.
SNPs extracted for chr2.
SNPs extracted for chr3.
SNPs extracted for chr4.
SNPs extracted for chr5.
SNPs extracted for chr6.
SNPs extracted for chr7.
SNPs extracted for chr8.
SNPs extracted for chr9.
SNPs extracted for chr10.
SNPs extracted for chr11.
SNPs extracted for chr12.
SNPs extracted for chr13.
SNPs extracted for chr14.
SNPs extracted for chr15.
SNPs extracted for chr16.
SNPs extracted for chr17.
SNPs extracted for chr18.
SNPs extracted for chr19.
SNPs extracted for chr20.
SNPs extracted for chr21.
SNPs extracted for chr22.
Merging SNPs extracted from each chromosome...
Created bed/bim/fam fileset with extracted SNPs: tmp_GENAL/4f4ce6a7_allchr
Extraction completed. 208(13.524%) SNPs were not extracted from the genetic data.
Computing a weighted PRS using tmp_GENAL/4f4ce6a7_allchr data.
The PRS computation was successful and used 1330/1538 (86.476%) SNPs.
PRS data saved to SBP_prs.csv
```

In our case, we have been able to find proxies for 571 of the 786 SNPs that were missing in the population genetic data (7 potential proxies have been removed because they were identical to SNPs already present in our data).

You can customize how the proxies are chosen with the following arguments:

- `reference_panel` : The reference population used to derive linkage disequilibrium values and find proxies. Defaults to `eur` .
- `kb` : Width of the genomic window to look for proxies (in thousands of base-pair positions). Defaults to `5000` .
- `r2` : Minimum linkage disequilibrium value with the original SNP for a proxy to be included. Defaults to `0.8` .
- `window_snps` : Width of the window to look for proxies (in number of SNPs). Defaults to `5000` .

> **Note:**
>
> You can call the `genal.Geno.prs` method on any `Geno` instance (containing at least the EA, BETA, and either SNP or CHR/POS columns). The data does not need to be clumped, and there is no limit to the number of instruments used to compute the scores.

#### Mendelian Randomization

To run MR, we need to load both our exposure and outcome SNP-level data in `genal.Geno` instances. In our case, the genetic instruments of the MR are the SNPs associated with blood pressure at genome-wide significant levels resulting from the clumping of the blood pressure GWAS. They are stored in our `SBP_clumped` `genal.Geno` instance which also include their association with the exposure trait (instrument-SBP estimates in the `BETA` column).

To get their association with the outcome trait (instrument-stroke estimates), we are going to use SNP-level data from a large GWAS of stroke performed by the GIGASTROKE consortium (https://www.nature.com/articles/s41586-022-05165-3):

```
stroke_gwas = pd.read_csv("GCST90104539_buildGRCh37.tsv",sep="\t")
```

We inspect it to determine the column names:

| chromosome | base_pair_location | effect_allele_frequency | beta | standard_error | p_value | odds_ratio | ci_lower |
|---|---|---|---|---|---|---|---|
| 5 | 29439275 | 0.3569 | 0.0030 | 0.0070 | 0.6658 | 1.003005 | 0.989337 |
| 5 | 85928892 | 0.0639 | -0.0152 | 0.0137 | 0.2686 | 0.984915 | 0.958820 |
| 10 | 128341232 | 0.4613 | 0.0025 | 0.0065 | 0.6998 | 1.002503 | 0.989812 |
| 3 | 62707519 | 0.0536 | 0.0152 | 0.0152 | 0.3177 | 1.015316 | 0.985514 |
| 2 | 80464120 | 0.9789 | 0.0057 | 0.0254 | 0.8223 | 1.005716 | 0.956874 |

We load it in a `genal.Geno` instance:

```
Stroke_Geno = genal.Geno(stroke_gwas, CHR = "chromosome", POS = "base_pair_location", EA =
"effect_allele", NEA = "other_allele", BETA = "beta", SE = "standard_error", P = "p_value", EAF =
"effect_allele_frequency", keep_columns = False)
```

We preprocess it as well to put it in the correct format and make sure there is no invalid values:

```
Stroke_Geno.preprocess_data(preprocessing = 'Fill_delete')
```

Now, we need to extract our instruments (SNPs of the `SBP_clumped` data) from the outcome data to obtain their association with the outcome trait (stroke). It can be done by calling the `genal.Geno.query_outcome` method:

```
SBP_clumped.query_outcome(Stroke_Geno, proxy = False)
```

Genal will print how many SNPs were successfully found and extracted from the outcome data:

```
Outcome data successfully loaded from 'b352e412' geno instance.
Identifying the exposure SNPs present in the outcome data...
1541 SNPs out of 1545 are present in the outcome data.
(Exposure data, Outcome data, Outcome name) stored in the .MR_data attribute.
```

Here as well you have the option to use proxies for the instruments that are not present in the outcome data:

```
SBP_clumped.query_outcome(Stroke_geno, proxy = True, reference_panel = "eur", kb = 5000, r2 = 0.6,
window_snps = 5000)
```

And genal will print the number of missing instruments which have been proxied:

```
Outcome data successfully loaded from 'b352e412' geno instance.
Identifying the exposure SNPs present in the outcome data...
1541 SNPs out of 1545 are present in the outcome data.
Searching proxies for 4 SNPs...
Using the EUR reference panel.
Found proxies for 4 SNPs.
(Exposure data, Outcome data, Outcome name) stored in the .MR_data attribute.
```

After extracting the instruments from the outcome data, the `SBP_clumped` `genal.Geno` instance contains an `MR_data` attribute containing the instruments-exposure and instruments-outcome associations necessary to run MR. Running MR is now as simple as calling the `genal.Geno.MR` method of the SBP_clumped `genal.Geno` instance:

```
SBP_clumped.MR(action = 2, exposure_name = "SBP", outcome_name = "Stroke_eur")
```

The `genal.Geno.MR` method prints a message specifying which SNPs have been excluded from the analysis (it depends on the action argument, as we will see):

```
Action = 2: 42 SNPs excluded for being palindromic with intermediate allele frequencies: rs11817866, rs3802517,
rs2788293, rs2274224, rs7310615, rs7953257, rs2024385, rs61912333, rs11632436, rs1012089, rs3851018, rs9899540,
rs4617956, rs773432, rs11585169, rs7796, rs2487904, rs12321, rs73029563, rs4673238, rs3845811, rs2160236, rs10165271,
rs9848170, rs2724535, rs6842486, rs4834792, rs990619, rs155364, rs480882, rs6875372, rs258951, rs1870735, rs1800795,
rs12700814, rs1821002, rs3021500, rs28601761, rs7463212, rs907183, rs534523, rs520015
```

It returns a dataframe containing the results for different MR methods:

| exposure | outcome | method | nSNP | b | se | pval |
|---|---|---|---|---|---|---|
| SBP | Stroke_eur | Inverse-Variance Weighted | 1499 | 0.023049 | 0.001061 | 1.382645e-104 |
| SBP | Stroke_eur | Inverse Variance Weighted (Fixed Effects) | 1499 | 0.023049 | 0.000754 | 4.390655e-205 |
| SBP | Stroke_eur | Weighted Median | 1499 | 0.022365 | 0.001337 | 8.863203e-63 |
| SBP | Stroke_eur | Simple mode | 1499 | 0.027125 | 0.007698 | 4.382993e-04 |
| SBP | Stroke_eur | MR Egger | 1499 | 0.027543 | 0.002849 | 1.723156e-21 |
| SBP | Stroke_eur | Egger Intercept | 1499 | -0.001381 | 0.000813 | 8.935529e-02 |

You can specify several arguments. We refer to the API for a full list, but the most important one is the `action` argument. It determines how palindromic SNPs are treated during the exposure-outcome harmonization step. Palindromic SNPs are SNPs where the nucleotide change reads the same forward and backward on complementary strands of DNA (for instance `EA = 'A'` and `NEA = 'T'`).

- `action = 1` : Palindromic SNPs are not treated (assumes all alleles are on the forward strand)
- `action = 2` : Uses effect allele frequencies to attempt to flip them (conservative, default)
- `action = 3` : Removes all palindromic SNPs (very conservative)

If you choose the option 2 or 3 (recommended), genal will print the list of palindromic SNPs that have been removed from the analysis.

By default, only some MR methods (inverse-variance weighted, weighted median, Simple mode, MR-Egger) are going to be run. But if you wish to run a different set of MR methods, you can pass a list of strings to the `methods` argument. The possible strings are:

- `IVW` for the classical Inverse-Variance Weighted method with random effects
- `IVW-RE` for the Inverse Variance Weighted method with Random Effects where the standard error is not corrected for under dispersion
- `IVW-FE` for the Inverse Variance Weighted with fixed effects
- `UWR` for the Unweighted Regression method
- `WM` for the Weighted Median method
- `WM-pen` for the penalised Weighted Median method
- `Simple-median` for the Simple Median method
- `Sign` for the Sign concordance test
- `Egger` for MR-Egger and the MR-Egger intercept
- `Egger-boot` for the bootstrapped version of MR-Egger and its intercept
- `Simple-mode` for the Simple mode method
- `Weighted-mode` for the Weighted mode method
- `all` to run all the above methods

For more fine-tuning, such as settings for the number of boostrapping iterations, please refer to the API.

Now, let's compare our python implementation of the MR methods with the widely used TwoSampleMR R package. First, let's check that the `action = 2` argument is excluding the same SNPs from the analysis. In R, we would have used:

```
dat <- harmonise_data(SBP_clumped, AS_outcome_glued, action = 2)
```

which outputs:

```
Harmonising exposure (qVf3Wu) and AS_EUR (Y65MM7)

Removing the following SNPs for being palindromic with intermediate allele frequencies:
rs1012089, rs10165271, rs11585169, rs11632436, rs11817866, rs12321, rs12700814, rs155364, rs1800795, rs1821002,
rs1870735, rs2024385, rs2160236, rs2274224, rs2487904, rs258951, rs2724535, rs2788293, rs28601761, rs3021500,
rs3802517, rs3845811, rs3851018, rs4617956, rs4673238, rs480882, rs4834792, rs520015, rs534523, rs61912333, rs6842486,
rs6875372, rs73029563, rs7310615, rs7463212, rs773432, rs7796, rs7953257, rs907183, rs9848170, rs9899540, rs990619
```

We verify that the same 42 SNPs are excluded (presented in a different order). Now, let's run all methods in python:

```
SBP_clumped.MR(action = 2, exposure_name = "SBP", outcome_name = "Stroke_eur", methods="all")
```

We obtain:

| exposure | outcome | method | nSNP | b | se | pval |
|---|---|---|---|---|---|---|
| SBP | Stroke_eur | Inverse-Variance Weighted | 1499 | 0.023049 | 0.001061 | 1.382645e-104 |
| SBP | Stroke_eur | Inverse Variance Weighted (Random Effects) | 1499 | 0.023049 | 0.001061 | 1.382645e-104 |
| SBP | Stroke_eur | Inverse Variance Weighted (Fixed Effects) | 1499 | 0.023049 | 0.000754 | 4.390655e-205 |
| SBP | Stroke_eur | Unweighted Regression | 1499 | 0.021233 | 0.073108 | 7.714853e-01 |
| SBP | Stroke_eur | Weighted Median | 1499 | 0.022365 | 0.001340 | 1.421514e-62 |
| SBP | Stroke_eur | Penalised Weighted Median | 1499 | 0.021216 | 0.001361 | 8.702012e-55 |
| SBP | Stroke_eur | Simple Median | 1499 | 0.021219 | 0.001284 | 2.582828e-61 |
| SBP | Stroke_eur | Sign concordance test | 1496 | 0.370321 | NaN | 1.928967e-47 |
| SBP | Stroke_eur | MR Egger | 1499 | 0.027543 | 0.002849 | 1.723156e-21 |
| SBP | Stroke_eur | Egger Intercept | 1499 | -0.001381 | 0.000813 | 8.935529e-02 |
| SBP | Stroke_eur | MR Egger bootstrap | 1499 | 0.030066 | 0.001966 | 0.000000e+00 |
| SBP | Stroke_eur | Egger Intercept bootstrap | 1499 | -0.002829 | 0.000687 | 0.000000e+00 |
| SBP | Stroke_eur | Simple mode | 1499 | 0.027125 | 0.007618 | 3.811199e-04 |
| SBP | Stroke_eur | Weighted mode | 1499 | 0.027125 | 0.006829 | 7.460982e-05 |

To use the same methods with the TwoSampleMR R package we run:

```
method_list = c("mr_ivw", "mr_ivw_fe",
                "mr_uwr",
                "mr_weighted_median",
                "mr_penalised_weighted_median",
                "mr_simple_median",
                "mr_sign",
                "mr_egger_regression",
                "mr_egger_regression_bootstrap",
               "mr_simple_mode",
               "mr_weighted_mode")
res <- mr(dat, method_list=method_list)
```

and we obtain:

| id.exposure | id.outcome | outcome | exposure | method | nsnp | b | se | pval |
|---|---|---|---|---|---|---|---|---|
| qVf3Wu | Y65MM7 | AS_EUR | exposure | Inverse variance weighted | 1499 | 0.02304861 | 0.001061261 | 1.382645e-104 |
| qVf3Wu | Y65MM7 | AS_EUR | exposure | Inverse variance weighted (fixed effects) | 1499 | 0.02304861 | 0.000754251 | 4.390655e-205 |
| qVf3Wu | Y65MM7 | AS_EUR | exposure | Unweighted regression | 1499 | 0.02123297 | 0.073108074 | 7.714853e-01 |
| qVf3Wu | Y65MM7 | AS_EUR | exposure | Weighted median | 1499 | 0.02236454 | 0.001365750 | 2.871914e-60 |
| qVf3Wu | Y65MM7 | AS_EUR | exposure | Penalised weighted median | 1499 | 0.02121585 | 0.001348878 | 9.643892e-56 |
| qVf3Wu | Y65MM7 | AS_EUR | exposure | Simple median | 1499 | 0.02121872 | 0.001286629 | 4.208342e-61 |
| qVf3Wu | Y65MM7 | AS_EUR | exposure | Sign concordance test | 1496 | 0.37032086 | NA | 1.928967e-47 |
| qVf3Wu | Y65MM7 | AS_EUR | exposure | MR Egger | 1499 | 0.02754335 | 0.002848883 | 1.723156e-21 |
| qVf3Wu | Y65MM7 | AS_EUR | exposure | MR Egger (bootstrap) | 1499 | 0.03008383 | 0.002157658 | 0.000000e+00 |
| qVf3Wu | Y65MM7 | AS_EUR | exposure | Simple mode | 1499 | 0.02712543 | 0.007397020 | 2.539175e-04 |
| qVf3Wu | Y65MM7 | AS_EUR | exposure | Weighted mode | 1499 | 0.02712543 | 0.006966826 | 1.031461e-04 |

We verify that the results match. The standard errors and p-values of the median, bootstrapped, and mode methods are not identical as they are calculated with generation of random data, which yields slightly different results at each run.

If you want to visualize the obtained MR results, you can use the `genal.Geno.MR_plot` method that will plot each SNP in an `effect_on_exposure x effect_on_outcome` plane as well as lines corresponding to different MR methods:

```
SBP_clumped.MR_plot(filename="MR_plot_SBP_AS")
```

MR plot You can select which MR methods you wish to plot with the `methods` argument. Note that for an MR method to be plotted, they must be included in the latest `genal.Geno.MR` call of this `genal.Geno` instance.

If you wish to include the heterogeneity values (Cochran's Q) in the results, you can use the `heterogeneity = True` argument in the `genal.Geno.MR` call. Here, the heterogeneity for the inverse-variance weighted method:

```
SBP_clumped.MR(action = 2, methods = ["Egger","IVW"], exposure_name = "SBP", outcome_name =
"Stroke_eur", heterogeneity = True)
```

And that will give:

| exposure | outcome | method | nSNP | b | se | pval | Q | Q_df | Q_pval |
|---|---|---|---|---|---|---|---|---|---|
| SBP | Stroke_eur | MR Egger | 1499 | 0.027543 | 0.002849 | 1.723156e-21 | 2959.965136 | 1497 | 1.253763e-98 |
| SBP | Stroke_eur | Egger Intercept | 1499 | -0.001381 | 0.000813 | 8.935529e-02 | 2959.965136 | 1497 | 1.253763e-98 |
| SBP | Stroke_eur | Inverse-Variance Weighted | 1499 | 0.023049 | 0.001061 | 1.382645e-104 | 2965.678836 | 1498 | 4.280737e-99 |

Here as well we can verify that the heterogeneity values are identical to those obtained in R:

```
mr_heterogeneity(dat)
```

| id.exposure | id.outcome | outcome | exposure | method | Q | Q_df | Q_pval |
|---|---|---|---|---|---|---|---|
| qVf3Wu | Y65MM7 | AS_EUR | exposure | MR Egger | 2959.965 | 1497 | 1.253763e-98 |
| qVf3Wu | Y65MM7 | AS_EUR | exposure | Inverse variance weighted | 2965.679 | 1498 | 4.280737e-99 |

As well as the Egger intercept value:

```
mr_pleiotropy_test(dat)
```

| id.exposure | id.outcome | outcome | exposure | egger_intercept | se | pval |
|---|---|---|---|---|---|---|
| qVf3Wu | Y65MM7 | AS_EUR | exposure | -0.001381477 | 0.0008126758 | 0.08935529 |

As expected, many MR methods indicate that SBP is strongly associated with stroke, but there could be concerns for horizontal pleiotropy (instruments influencing the outcome through a different pathway than the one used as exposure) given the almost significant MR-Egger intercept p-value. To investigate horizontal pleiotropy in more details, a very useful method is Mendelian Randomization Pleiotropy RESidual Sum and Outlier (MR-PRESSO). MR-PRESSO is a method designed to detect and correct for horizontal pleiotropy. It will identify which instruments are likely to be pleiotropic on their effect on the outcome, and it will rerun an inverse-variance weighted MR after excluding them. It can be run using the `genal.Geno.MRpresso` method:

```
SBP_clumped.MRpresso(action = 2, n_iterations = 30000)
```

As with the `genal.Geno.MR` method, the `action` argument determines how the pleiotropic SNPs will be treated. The output is a list containing:

- A table containing the original and outlier-corrected inverse variance-weighted results.
- The global test p-value indicating the presence of horizontal pleiotropy.
- A dataframe of p-values, one for each instrument, representing the likelihood that this instrument is pleiotropic (only relevant if the global test is significant).
- A dictionary containing the outputs of the distortion test. This test assesses whether the removal of the pleiotropic instruments has significantly altered the original MR estimate.
    - An array containing the indices of the pleiotropic SNPs.
    - The coefficient of the distortion test.
    - The p-value of the distortion test.

| exposure | method | nSNP | b | se | pval |
|---|---|---|---|---|---|
| BETA_e | Raw | 1499 | 0.023051 | 0.001061 | 3.674047e-91 |
| BETA_e | Outlier-corrected | 1487 | 0.022926 | 0.001028 | 2.860803e-95 |

```
{'RSSobs': 2970.590771018229, 'Global_test_p': '< 3.3e-05'}
```

Here again, we can compare our results to those obtained in R, with the original MR-PRESSO package:

```
run_mr_presso(dat, NbDistribution = 30000)
```

| Exposure | MR Analysis | Causal Estimate | Sd | T-stat | P-value |
|---|---|---|---|---|---|
| beta.exposure | Raw | 0.02304861 | 0.001061261 | 21.71814 | 3.801497e-91 |
| beta.exposure | Outlier-corrected | 0.02310329 | 0.001035207 | 22.31755 | 2.065088e-95 |

```
[[1]]$`MR-PRESSO results`$`Global Test`$RSSobs
[1] 2970.515

[[1]]$`MR-PRESSO results`$`Global Test`$Pvalue
[1] "<3.33333333333333e-05"
```

MR-PRESSO is based on the generation of random data to identify outliers, which leads to results slightly varying between runs. Genal implements MR-PRESSO in parallel, allowing it to take advantage of all available CPU cores. This enables Genal to execute MR-PRESSO much faster than the original R version. For instance, in this example involving 1499 SNPs and 30,000 iterations of random data generation, the original R version required 15 hours and 50 minutes, whereas our Genal version, running on an AMD Ryzen 5900x CPU with 12 cores at 3.7 GHz, completed the task in 1 hour and 58 minutes. This is a 87.5% reduction in execution time.

#### SNP-association testing

We may want to calibrate instrument-trait estimates in a specific population for which we have individual-level data (genetic files as well as phenotypic data). For instance, if the GWAS of SBP was done in a european population, we may want to adjust the estimates based on data coming from a population of a different ancestry. This can be done in 2 steps:

- Loading the phenotypic data in a dataframe and calling the `genal.Geno.set_phenotype` method
- Calling the `genal.Geno.association_test` method to run the association tests and update the estimates Let's start by loading phenotypic data::

```
df_pheno = pd.read_csv("path/to/trait/data")
```

> **Note:**
>
> One important detail is to make sure that the individual IDs are identical between the phenotypic data and the genetic data for the target population.

Then, it is advised to make a copy of the `genal.Geno` instance containing our instruments as we are going to update their coefficients and to avoid any confusion:

```
SBP_adjusted = SBP_clumped.copy()
```

We can then call the `genal.Geno.set_phenotype` method, specifying which column contains our trait of interest (for the association testing) and which column contains the individual IDs:

```
SBP_adjusted.set_phenotype(df_pheno, PHENO = "htn", IID = "IID")
```

At this point, genal will identify if the phenotype is binary or quantitative in order to choose the appropriate regression model. If the phenotype is binary, it will assume that the most frequent value is coding for control (and the other value for case), this can be changed with `alternate_control = True` :

```
Detected a binary phenotype in the 'PHENO' column. Specify 'PHENO_type="quant"' if this is incorrect.
Identified 0 as the control code in 'PHENO'. Set 'alternate_control=True' to inverse this interpretation.
The phenotype data is stored in the .phenotype attribute.
```

We can then run the association tests, specifying the path to the genetic files in plink format, and any columns we may want to include as covariates in the regression tests:

```
SBP_adjusted.association_test(covar=["age"], path = "path/to/genetic/files")
```

Genal will print information regarding the number of individuals used in the tests and the kind of tests performed. It is advised to make sure that these information are consistent with your data:

```
CHR/POS columns present: SNPs searched based on genomic positions.
Extracting SNPs for each chromosome...
SNPs extracted for chr1.
SNPs extracted for chr2.
SNPs extracted for chr3.
SNPs extracted for chr4.
SNPs extracted for chr5.
SNPs extracted for chr6.
SNPs extracted for chr7.
SNPs extracted for chr8.
SNPs extracted for chr9.
SNPs extracted for chr10.
SNPs extracted for chr11.
SNPs extracted for chr12.
SNPs extracted for chr13.
SNPs extracted for chr14.
SNPs extracted for chr15.
SNPs extracted for chr16.
SNPs extracted for chr17.
SNPs extracted for chr18.
SNPs extracted for chr19.
SNPs extracted for chr20.
SNPs extracted for chr21.
SNPs extracted for chr22.
Merging SNPs extracted from each chromosome...
Created bed/bim/fam fileset with extracted SNPs: tmp_GENAL/e415aab3_allchr
39131 individuals are present in the genetic data and have a valid phenotype trait.
Running single-SNP logistic regression tests on tmp_GENAL/e415aab3_allchr data with adjustment for: ['age'].
The BETA, SE, P columns of the .data attribute have been updated.
```

The `BETA`, `SE`, and `P` columns of the `SBP_adjusted.data` attribute have been updated with the results of the association tests.

##### Lifting

It is sometimes necessary to lift the SNP data to a different build. For instance, if the genetic data of our target population is in build 38 (hg38), but the GWAS summary statistics are in build 37 (hg19). This can easily be done in genal using the `genal.Geno.lift` method:

```
SBP_clumped.lift(start = "hg19", end = "hg38", replace = False)
```

This outputs a table with the lifted SBP instruments (stored in the `SBP_clumped` instance) from build 37 (hg19) to build 38 (hg38). We specified `replace = False` to not modify the `SBP_clumped.data` attribute, but we may want to modify it (before running a PRS in a population stored in build 38 for instance). Genal will download the appropriate chain files required for the lift, and it will be done in python by default. However, if you plan to lift large datasets of SNPs (the whole summary statistics for instance), it may be useful to install the LiftOver executable that will run faster than the python version. It can be downloaded here: https://genome-store.ucsc.edu/ You will need to create an account, scroll down to "LiftOver program", add it to your cart, and declare that you are a non-profit user.

You can specify the path of the LiftOver executable to the `liftover_path` argument:

```
SBP_Geno.lift(start = "hg19", end = "hg38", replace = False, liftover_path = "path/to/liftover/exec")
```

#### Examples from published papers

##### Polygenic Susceptibility to Hypertension and Cognitive Performance

This paper ([https://pubmed.ncbi.nlm.nih.gov/37295956/]) reports that higher polygenic susceptibility to high blood pressure is associated with worse cognitive performance in middle aged adults. Polygenic susceptibility to high blood pressure was modeled using two polygenic risk scores for systolic or diastolic blood pressure. The SNPs and their associations with systolic and diastolic blood pressure are provided in Supplementary Table 1:

| CHR | BP | SNP | A1 | A2 | Beta_SBP | se_SBP | Beta_DBP | se_DBP |
|---|---|---|---|---|---|---|---|---|
| 1 | 1687482 | rs2076328 | T | G | -0.2824 | 0.0498 | -0.0965 | 0.0292 |
| 1 | 2187085 | rs260508 | T | G | 0.1297 | 0.0441 | 0.0854 | 0.0323 |
| 1 | 3328659 | rs2493292 | T | C | 0.2686 | 0.0680 | 0.1674 | 0.0401 |
| 1 | 6278414 | rs709209 | A | G | 0.0935 | 0.0528 | -0.0271 | 0.0308 |
| 1 | 7739250 | rs4908678 | T | C | -0.1203 | 0.0484 | -0.1065 | 0.0285 |

Producing the polygenic risk scores in the UK Biobank from the table of SNP information can be done in just a few lines with genal:

```
## Load libraries and paths
import pandas as pd
import genal
genal.set_plink(path="path/to/plink1.9") # Set the path to plink 1.9
ukb_geno_path = "path/to/UKB_geno_files/plink_$" #Path to the UKB plink files broken down by
chromosomes

## Build the PRS for systolic blood pressure
df_bp = pd.read_csv("BP_SNPs.csv") # Load the table with the SNP information
SBP_Geno = genal.Geno(df_bp, CHR = "CHR", POS = "BP", EA = "A1", NEA = "A2", BETA = "Beta_SBP", SE =
"se_SBP", SNP = "SNP", keep_columns = False) #Declare a Geno object for the systolic blood pressure
information
SBP_Geno.preprocess_data(preprocessing = 'Fill_delete', reference_panel="eur") #Preprocess it (not
necessary in this case as the data is already cleaned)
SBP_Geno.prs(name = "SBP_prs", path = ukb_geno_path) #Compute the PRS for systolic blood pressure

## The steps are similar to build the PRS for diastolic blood pressure
DBP_Geno = genal.Geno(df_bp, CHR = "CHR", POS = "BP", EA = "A1", NEA = "A2", BETA = "Beta_DBP", SE =
"se_DBP", SNP = "SNP", keep_columns = False)
DBP_Geno.preprocess_data(preprocessing = 'Fill_delete', reference_panel="eur")
DBP_Geno.prs(name = "DBP_prs", path = ukb_geno_path)
```

The two risk scores for systolic and diastolic blood pressure are not stored in .csv files and can easily be used for further analyses.

##### Association of Poor Oral Health With Neuroimaging Markers of White Matter Injury

This paper ([https://pubmed.ncbi.nlm.nih.gov/38165331/]) investigates whether poor oral health associates with worse neuroimaging markers of white matter injury. A robust way to study such a relationship is to combine both observational (such as cross-sectional or longitudinal design) and Mendelian Randomization (MR) analyses. MR is very useful to confirm observational findings (as it is less prone to confounding) and it also allows to assess the causality of associations with a higher level of confidence than observational studies alone. This is the framework that was used in this study, and we present here how genal makes the MR part of the analysis straighforward. We first load libraries and paths, and create a `genal.Geno` instance with the summary statistics of the GWAS of poor oral health (which uses a composite trait of caries, dentures, and filled or missing teeth). We clump the summary statistics to select our genetic instruments (independent variants strongly associated with our exposure trait, poor oral health).:

```
## Load libraries and paths
import pandas as pd
import genal
genal.set_plink(path="path/to/plink1.9") # Set the path to plink 1.9
ukb_geno_path = "path/to/UKB_geno_files/plink_$" # Path to the UKB plink files broken down by chromosomes

## Obtain instruments
df_OH=pd.read_csv("DMFS_Dentures.txt",sep="\t") # Load the GWAS summary statistics of poor oral health (OH)
OH_Geno = genal.Geno(df_OH, SNP="rsid", CHR="CHR", POS="POS", BETA="beta", SE="se", EA="Allele1", NEA="Allele2", EAF="Freq1", P="p", keep_columns = False) # Create a Geno object with the summary statistics
OH_Geno.preprocess_data(preprocessing = 'Fill_delete', reference_panel="eur") # Preprocess the summary statistics to format the data and remove erroneous information
OH_Instruments = OH_Geno.clump(p1=5e-8,r2=0.1,kb=250, reference_panel = "eur") # Clump the summary statistics to obtain the instruments
```

Now that we have an `OH_clumped` object containing our instruments and their association statistics with the exposure, we will run association tests to determine the association statistics of these instruments in regard to our outcomes (3 neuroimaging markers: white matter hyperintensities (WMH), fractional anisotropy (FA), and mean diffusivity (MD)). We will then run one MR analysis for each outcome. All of these steps can be performed in a loop while storing the MR results in a dictionary:

```
## Load the dataframe containing the individual-level data for our outcomes
df_phenotypes=pd.read_csv("/path/to/ukb_mri_dti.csv")

## Run the association tests and MR for each outcome in a loop with results stored in a dictionary
MR_results = {} # Dictionary to store MR results
for outcome in ("PC1_fa","PC1_md","wmh"): # Iterate over every outcome of interest
    Outcome_Instruments = OH_Instruments.copy() # Create a copy of the instruments
    Outcome_Instruments.set_phenotype(df_phenotypes,PHENO=outcome,IID="IID") # Attach the phenotype dataset to the instruments copy and specify the outcome name
    Outcome_Instruments.association_test(covar = ["age", "sex", "PC_1", "PC_2", "PC_3", "PC_4"],path = ukb_geno_path) # Update the coefficients of the instruments copy to reflect their associations with the outcome
    OH_Instruments.query_outcome(OH_Instruments_outcome, proxy = False) # Query the instruments copy (now containing associations with the outcome)
    MR_results[outcome] = OH_clumped.MR(action = 1, exposure_name = "Oral health", outcome_name = outcome, nboot=1000) # Run the MR and store it in the dictionary
```
